# Population-Level Heart Attack Risk Assessment Using Self-Reported Data: A Cross-Sectional Study with BRFSS 2023

**DOI:** 10.1101/2025.07.02.25330700

**Authors:** Adam Schomer, Stephen Schomer

## Abstract

The major cause of mortality in the country is still myocardial infarction, which is shaped by demographic, socioeconomic, and clinical elements. Despite research, there are still major gaps in understanding how these elements affect different populations. The present prediction models rely on laboratory values, which may be limited in resource-scarce settings. This study aimed to create and evaluate a heart attack risk prediction model by using a staging technique with demographic and clinical risk factors from the Behavioral Risk Factor Surveillance System 2023. Starting with a sample of 430,755 participants, this cross-sectional study randomly selected 100,000 participants with a fixed seed code cleaning the missing data and subsequently performing multiple imputations. The model adopted a staging approach, with four stages: (1) demographic factors including age, sex, race, income, and education; (2) diabetes (3) hypertension, and (4) cholesterol. The model performance was evaluated using AUC-ROC, calibration plots, and a sensitivity analysis. The overall heart attack prevalence based on these results was 5.3% (95% CI: 5.0-5.5%). The final model showed strong discrimination (AUC-ROC=0.80) and strong calibration. The strongest risk factors in this predictive model were ages ≥ 70 (OR=6.58, 95% CI: 5:37-8.06), high cholesterol (OR=1.77), hypertension (OR=1.81), and diabetes (OR=1.50). There were protective associations identified, including female sex (OR=0.49) and having a college education (OR=0.42). Black (OR=0.62, 95% CI: 0.50-0.76), Hispanic (OR=0.76, 95% CI: 0.58-0.99), and Native American/Alaskan (OR=0.66, 95% CI: 0.54-0.81) people had lower odds of self-reported heart attack following adjustment compared to White participants. The model performance was consistent across sex groups, with an AUC-ROC, males=0.794 and females=0.787. The parsimonious model illustrated strong prediction can be achieved using self-reported data using a staging approach, which highlights incremental value of each risk factor contributing to a population-level heart attack risk assessment.

## Introduction

With heart attacks being the lens of focus in this study, cardiovascular disease (CVD) remains one of the leading causes of death in the country. With about 800,000 heart attacks per year, heart attacks remain a significant issue, which has scientists exploring new ways to lower its occurrence and enhance its detection[1]. The American Heart Association (AHA) and Center for Disease Control and Prevention (CDC) mentions how heart attacks result in about 2,500 deaths per day[2][3]. This has been supported by the CDC, which went even further to say every 33 seconds, someone dies from CVD[3]. The prevalence of heart disease is nothing short of alarming and warrants further attention.

Heart disease is complicated, and the development of the disease can be complex, as it integrates various interconnected elements. These interconnected elements are largely influenced by various social determinants of health (SDoH), including demographics, socioeconomic status, lifestyle habits, behavioral influences, and cardiometabolic comorbidities[4][5]. These key factors play a role in the development of a heart attack, such as the patients age, sex, smoking status, marital status, physical activity levels, and clinical comorbidities[6][7]. These factors have been largely associated with the development of a heart attack and have been thoroughly researched to better understand how this impacts patients and public health[8].

Though years of research have focused on understanding heart attacks, including assessing different risk factors and treatments, heart attacks continue to persist[9]. The outcome of heart attacks remains constant across the research efforts, indicating that different factors influence their development, including socioeconomic and racial disparities[7][10]. These disparities are also linked to higher rates of CVD disease risk factors, including hypertension, diabetes, hyperlipidemia, and kidney disease[11][12][13].

Despite extensive decades of research, there are still significant gaps in understanding how these key risk factors impact the diverse populations across the nation. The purpose of this study is to address the gap in knowledge by exploring these interconnected factors using Behavioral Risk Factor Surveillance System (BRFSS) from 2023, which provides comprehensive population-level data. Therefore, this study aims to answer the following questions:

1. Which populations have the highest heart attack burden?
2. Are there significant health disparities in heart attack occurrence?
3. Which factors are most strongly associated with heart attacks?
4. Which associations remain significant after accounting for other factors?

This study hypothesizes that demographic variables including age, sex, socioeconomic status, and education would establish a foundation discrimination for heart attack risk, with the sequential addition of clinical factors (diabetes, hypertension, and hyperlipidemia), which would yield incremental improvements in prediction using a staging approach.

## Methodology

### 2.1 Data Source and Study Population

The dataset that was analyzed was from the BRFSS 2023 landline and cellphone survey, a comprehensive national survey that gathers information on a wealth of variables. The data is self-reported on a survey, which is important to note. The dataset analyzed in this study contained 430,755 participants, which were adults in the United States, ages 18 and older. In effort to strengthen the results of the data, 100,000 participants were randomly selected using R- studio, which was about 23.2% of the participants, for the analysis. The sample information can be found on Table 1. The participants who had missing information on their heart attack status, n=2,568, were excluded from the randomized sampling because it only accounted for 0.6% of the sample.

**Table 1.** Demographic, socioeconomic, and clinical characteristics of the study population.

| Characteristic | Heart Attack Status |  |  | p-value<sup>2</sup> |
| --- | --- | --- | --- | --- |
|  | No<sup>1</sup> | Yes<sup>1</sup> | N = 5,380<sup>1</sup> |  |
| **Sex** |  |  |  | <0.001 |
| Male | 46,740 (47%) | 43,471 (46%) | 3,269 (61%) |  |
| Female | 53,260 (53%) | 51,149 (54%) | 2,111 (39%) |  |
| **Age Group** |  |  |  | <0.001 |
| 18-49 years | 36,316 (36%) | 35,929 (38%) | 387 (7.2%) |  |
| 50-69 years | 36,288 (36%) | 34,253 (36%) | 2,035 (38%) |  |
| ≥70 years | 27,396 (27%) | 24,438 (26%) | 2,958 (55%) |  |
| **Race/Ethnicity** |  |  |  | <0.001 |
| White | 73,808 (74%) | 69,518 (73%) | 4,290 (80%) |  |
| Black | 7,875 (7.9%) | 7,521 (7.9%) | 354 (6.6%) |  |
| Hispanic | 5,672 (5.7%) | 5,444 (5.8%) | 228 (4.2%) |  |
| Asian | 2,499 (2.5%) | 2,341 (2.5%) | 158 (2.9%) |  |
| Native American/Alaskan | 10,146 (10%) | 9,796 (10%) | 350 (6.5%) |  |
| **Income** |  |  |  | <0.001 |
| \$25k-<35k | 11,385 (11%) | 10,564 (11%) | 821 (15%) | |
| \$35k-<50k | 13,797 (14%) | 12,973 (14%) | 824 (15%) | |
| \$50k-<100k | 52,216 (52%) | 50,056 (53%) | 2,160 (40%) | |
| <\$25k | 15,178 (15%) | 13,788 (15%) | 1,390 (26%) | |
| ≥\$100k | 7,424 (7.4%) | 7,239 (7.7%) | 185 (3.4%) | |
| **Education** |  |  |  | <0.001 |
| Less than high school | 5,784 (5.8%) | 5,238 (5.5%) | 546 (10%) |  |
| High school | 24,799 (25%) | 23,197 (25%) | 1,602 (30%) |  |
| Some college | 26,406 (26%) | 24,796 (26%) | 1,610 (30%) |  |

| Heart Attack Status |  |  |  |  |
| --- | --- | --- | --- | --- |
| Characteristic | No<sup>1</sup> | Yes<sup>1</sup> | N = 5,380<sup>1</sup> | p-value<sup>2</sup> |
| College graduate | 43,011 (43%) | 41,389 (44%) | 1,622 (30%) |  |
| **Diabetes Status** |  |  |  | <0.001 |
| No | 2,342 (2.3%) | 2,158 (2.3%) | 184 (3.4%) |  |
| Pre-diabetes | 83,249 (83%) | 79,907 (84%) | 3,342 (62%) |  |
| Yes | 14,409 (14%) | 12,555 (13%) | 1,854 (34%) |  |
| **Hypertension Status** |  |  |  | <0.001 |
| Borderline | 1,005 (1.0%) | 969 (1.0%) | 36 (0.7%) |  |
| No | 57,569 (58%) | 56,195 (59%) | 1,374 (26%) |  |
| Yes | 41,426 (41%) | 37,456 (40%) | 3,970 (74%) |  |
| **High Cholesterol** |  |  |  | <0.001 |
| No | 59,607 (60%) | 57,783 (61%) | 1,824 (34%) |  |
| Yes | 40,393 (40%) | 36,837 (39%) | 3,556 (66%) |  |

Note: Analysis was based on a random sample of 100,000 BRFSS participants from the original sample size. The missing data was most common in income (20.1%) and cholesterol status (12.6%). P-values are from Pearson's Chi-square Test.
<sup>1</sup>n (%): N = 5,380 refers to the number of participants who reported having a heart attack (5.3% of the total sample)
<sup>2</sup>Pearson's Chi-squared test

### 2.2 Outcome Measure

The primary outcome focused on the participants who self-reported having a history of a heart attack, and the question that was asked to the participants was “has a doctor, nurse, or other health professional ever told you that you had a heart attack, also called a myocardial infarction?" The responses were binary, sorted into “yes” and “no” and any answer with “Don’t know/Not sure” or “refused” were treated as missing data and was excluded from the analysis (which was n=2,568 or 0.6% of the sample).

### 2.3 Variables and Definitions

There were several predictor variables included in the study to adjust for confounding.

#### Stage 1

Demographic characteristics: The three demographic characteristics are age, sex, and race. The age predictor variables were stratified into three categories, (under 50 years old [reference], 50-69 years old, and 70 years and older). The sex characteristic focused on male (reference) and female. The last characteristic is race, which evaluated White (reference), Black, Hispanic, Asian, and Native American/Alaskan. Socioeconomic characteristics: there are three socioeconomic characteristics included in this analysis, which are income, education, and employment status. The income predictor variable was stratified into five categories, starting with under $25k per year, then $25-35k per year, then $35-50k per year, $50k-100k per year, and then 100k+ per year (100k+ is the reference). The next predictor variable is education, which included less than high school, high school, some college, and college graduate (this is the reference).

#### Stage 2: Diabetes Status

The next clinical predictor variable is diabetes status, stratified into four categories, starting with No (reference group), pre-diabetes/borderline, or yes

#### Stage 3: Hypertension Status

The Next predictor variable focuses on hypertensive status, which is stratified into four categories, starting with No (reference group), then borderline, and yes

#### Stage 4: Cholesterol Status

The last clinical predictor variable is hypercholesterolemia, which is set to no (reference) and yes.

### 2.4 Statistical Analysis

The statistical testing in this analysis focused on logistic regression to understand and predict the association between heart attack risks (outcome variable) and the predictor variables listed above. The model analyzed both crude (unadjusted) and adjusted odds ratios with 95% CI to have a better understanding of these associations. The final model was a multivariable logistic regression using all predictor variables, which allowed this analysis to adjust for confounding.

The statistical analysis also aimed to quantify the magnitude of these confounding variables, therefore, the percentage difference between the crude and adjusted odds ratios was analyzed as well.

#### 2.4.1 Missing Data Handling

The missing data was assessed for all variables to provide an accurate assessment of the research questions. The analysis used multiple imputations to address the missing data, specifically Multiple Imputation using Chained Equations (MICE), which was performed. The multiple imputations used 5 imputations with 3 iterations using the MICE package in R-studio. The imputation model included all variables that were used in the analysis, plus the survey designed variables. Convergence was assessed through the use of diagnostic plots, and all analyses were conducted using the datasets that were imputed, and the combined results used Rubins rules to improve the accuracy of the results. Convergence of the MICE algorithm was confirmed through:

- **Trace plots** showing stochastic stability across 3 iterations
- **Gelman-Rubin statistics** <1.1 for all imputed variables
- **Fraction of Missing Information (FMI)** below 0.3 for key clinical variables (hypertension=0.18, cholesterol=0.25)

#### 2.4.2 Survey Design Considerations

BRFSS is a complex survey that uses stratification, clustering, and weights, all which was accounted for in this analysis. The BRFSS data collected had survey weights attached to them to generate nationally representative estimates to make the results more generalizable. The survey analysis conducted used the package “survey” from R-studio, and used the option ‘survey.lonely.psu’ and it was set to “adjust” to handle singleton primary sampling units.

#### 2.4.3 Staged Modeling Approach

The staged modeling approach followed 4 stages, which were added sequentially as predictor variables.

- Stage 1: Demographic factors only
- Stage 2: Stage 1 + diabetes status
- Stage 3: Stage 2 + hypertension status
- Stage 4: Stage 3 + cholesterol status

Before each stage was added, the research fit survey-weighted logistic regression models, predicting heart attack status. The results were pooled together from the imputed datasets using Rubin’s Rules and the Odds Ratios were set with a 95% confidence interval, and all the p-values were also calculated for the predictor variables. This staged approach was selected to systematically evaluate the incremental value of adding modifiable clinical risk factors to baseline demographic and socioeconomic characteristics. The sequence progresses from non-modifiable factors (demographics) to increasingly modifiable clinical factors, allowing us to assess which variables provide the most significant improvements in predictive performance.

#### 2.4.4 Model Performance Assessment

The model performance was assessed as well using the Area Under the Receiver Operating Characteristic Curve (AUC-ROC), to have a full picture of these statistical findings. The model calibration test was also performed using a calibration slope and interception. All statistical analyses were performed using R-studio 4.4.3 with a level of significance with the p < 0.05. Calibration was also used for evaluation, using calibration plots and comparing the predicted probabilities to the observed outcomes. The model also quantified the calibration by using the Integrated Calibration Index.

#### 2.4.5 Sensitivity analysis

The sensitivity analysis was conducted per each stage to assess the models performance across each stage and their key subgroups.

- Sex-stratified analysis: the model performance was evaluated separated for males and females
- Age-stratified analysis: the model performance was evaluated across the three age groups that were selected, such as 18-49, 50-69, and ≥70 years
- Variable Importance Analysis: The last sensitivity analysis was used to evaluate model performance with sequential removal of individual variables to quantify the variables and their contribution to model discrimination.

#### 2.4.6 Software and Reproducibility

The analysis was conducted in R-studio, using R-studio 4.4.3 and it included packages such as "haven", "survey", "dplyr", "ggplot2", "knitr", "kableExtra", "janitor", "tidyr", "scales", "viridis", "gridExtra", "usmap", "forcats", "patchwork", "gtsummary", "mice", "mitools", "car", "parallel", "pROC.” The complete analytical code is available at https://github.com/AdamSchomer/HeartAttackRisk

#### 2.4.7 Statistical Power

A post hoc power analysis was conducted using G*Power 3.1.9.7 software. Although our final sample size was 430,755 participants, there were 100,000 participants randomized using a fixed seed (set.seed()) for reproducibility. The purpose of this was to make sure the subsample was reproducible and kept its statistical properties. The sample achieved 100% power to detect an odds ratio of 1.5 at α=0.05 (two-tailed) with the assumption of a baseline event probability of 0.2. This indicates the study was adequately powered to detect even relatively small effects.

### 2.5 Ethics Oversight and Compliance

This secondary analysis used de-identified BRFSS 2023 data and was conducted under the CDC’s ongoing Institutional Review Board exemption (Protocol #0920-1061). This falls under 45 CFR §46.104(d)(4)(i) for analysis of publicly available, de-identified surveillance data. The participants in this BRFSS survey provided written informed consent during the original data collection aspect in accordance to federal survey protocols. As independent researchers, there were various measures that were taken to ensure and uphold ethical compliance:

- Privacy Protection: The authors made no re-identification attempts when using BRFSS data, and this research analyzed pre-collected records without direct identifiers.
- Regulatory Compliance: The study procedures align with the Declaration of Helsinki and Belmont Report principles, which emphasized on the importance of beneficence through focus on cardiovascular risk equity improvements.
- Transparency: The complete analytical code is available at https://github.com/AdamSchomer/HeartAttackRisk which contains no protected health

information and adheres to BRFSS data use agreements.

- BRFSS 2023 data can be publicly accessed, as no restrictions apply to accessing this data via CDC portal: https://www.cdc.gov/brfss/annual_data/annual_2023.html

## Results

### 3.1 Study Population Characteristics

The study focused on 430,755 participants who participated in the BRFSS national survey and was narrowed down to 100,000 participants that were randomly selected using R- studio. The 100,000-participant sample was designed to provide information on the overall prevalence of self-reported heart attacks, which was 5.3% (95% CI: 5.0-5.5%). The baseline characteristics of this study were stratified by heart attack status, which can be seen in Table 1. The missing data was noted to be common in income at 20.1% and cholesterol status at 12.6%, with other variables being lower rates. Table 1 displays the distribution of demographic, socioeconomic, and clinical characteristics, which was stratified by heart attack status. The table highlights notable differences between participants with and without self-reported heart attacks across all examined variables.

### 3.2 Which populations Have the Highest Heart Attack Burden?

The prevalence of heart attack varied significantly across the demographic groups as seen in table 1, where prevalence increased markedly with age: 1.0% (95% CI: 0.8-1.1%) among adults between the ages of 18 and 49, 5.7% (95% CI: 5.3-6.2%) among adults between the ages of 50 and 69, and 10.9% (95% CI: 10.2-11.6%) among adults from the ages of 70 years and over. The prevalence pattern for men were significantly higher than women, with men having 6.7%, with a 95% CI: 6.3-7.1% and woman having 3.9% prevalence, with 95% CI: 3.6-4.2%, and a p<0.001.

Other prevalence patterns emerged as well, with strong socioeconomic gradients being observed and a noteworthy decrease in prevalence as education and income increased. Adults who had less than a high school education experienced the highest prevalence (10.2%, 95% CI: 8.8-11.6%), which was compared to college graduates who had a significantly lower prevalence (3.6%, 95% CI: 3.3-4.0% p<0.001). The prevalence followed a similar pattern in income, where the highest prevalence was with those who made less than $25,000 per year (8.8%, 95% CI: 7.9- 9.8%), which compared to participants who made $100,000 or more annually (2.6%, 95% CI: 1.8-3.5%, p<0.001).

Clinical factors were also following similar patterns, where participants diagnosed with diabetes, hypertension, and high cholesterol experienced significantly higher prevalence patterns compared to participants who did not have these conditions. Diabetes prevalence was noted to be (12.9%, 95% CI: 11.9-13.9%), while hypertension was (9.6%, 95% CI: 9.0-10.1%), and lastly high cholesterol (8.8%, 95% CI: 8.3-9.3%), all with a p<0.001.

### 3.3 Health Disparities in Heart Attack Occurrence

There were significant disparities demonstrated across multiple demographic and socioeconomic variables with regards to heart attack occurrence. The statistical testing performed in this research confirmed there were significant differences in heart attack prevalence across all the characteristics evaluated (p<0.001 for all ages). The age-related disparities were noteworthy, which showed that adults ≥70 years experiencing heart attacks at nearly 11 times the rate of those aged 18-49 years (10.9% vs. 1.0%). The gender gap was similarly notable, with men experiencing heart attacks at a rate 72% higher than women (6.7% vs 3.9%).

The socioeconomic disparities were also noteworthy and showed a clear gradient. The educational attainment showed an inverse relationship with heart attack prevalence, where the rates were nearly three times higher in participants who had less than a high school education when compared to college graduates (10.2% vs 3.6%). The income variable demonstrated disparities as well, which followed a similar pattern with heart attack prevalence, showing over three times higher likelihood in the lowest income category (<$25k annually) when compared to the highest income category (≥$100k) (8.8% vs. 2.6%).

The racial and ethnic disparities that emerged were evident in the unadjusted prevalence figures, with White participants showing the highest prevalence (5.9%) compared to Black participants (4.4%), Native American/Alaskan participants (3.7%), and Hispanic participants (3.3%). However, the adjusted odds ratios demonstrated lower risk among these groups after controlling for socioeconomic, demographic, and clinical comorbidities, which potentially suggest there may be reporting or access disparities, rather than true biological protective factors.

The clinical comorbidity disparities were significant, with heart attack prevalence being considerably higher in participants who reported diabetes (12.9% vs 7.3% without), hypertension (9.6% vs. 2.3% without), and high cholesterol (8.8% vs. 2.8% without). These disparities underpin the various cardiovascular risk factors in vulnerable populations. Figure 1 demonstrates that prevalence differences across key demographic and clinical factors, which highlights the significant disparities observed in this study.

**Fig 1.**
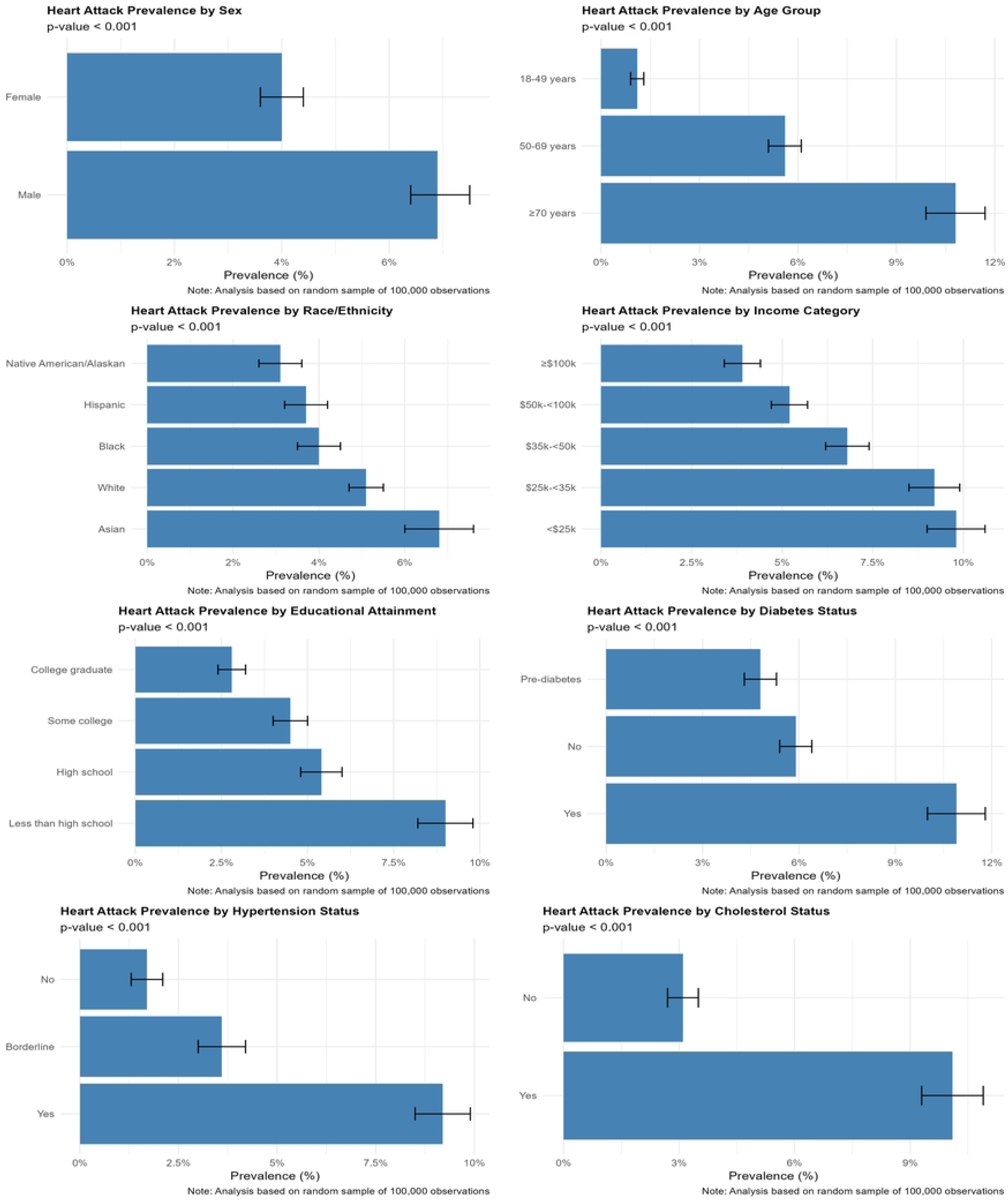
Heart Attack Prevalence by Key Demographic and Clinical Factors. This figure displays heart attack prevalence (percentage with 95% confidence intervals) across demographic and clinical subgroups. The horizontal bars represent prevalence estimates, with longer bars indicating higher heart attack burden. The error bars represent 95% confidence intervals, with non-overlapping intervals indicating statistically significant differences between groups (p<0.001 for all comparisons)

### 3.4 Factors Most Strongly Associated with Heart Attacks

The risk factors in the fully adjusted model, Stage 4 Model (Table 2, Figure 2), demonstrated that age was consistently a strong predictor of heart attack risk, with adults aged 50-69 years old (OR=3.82, 95% CI, 3.11-4.68) and adults 70-years and older (OR=6.58, 95% CI: 5.37-8.06) were at substantially higher risk compared to those ages 18-49 years old (p<0.001 for both groups).

**Fig 2.**
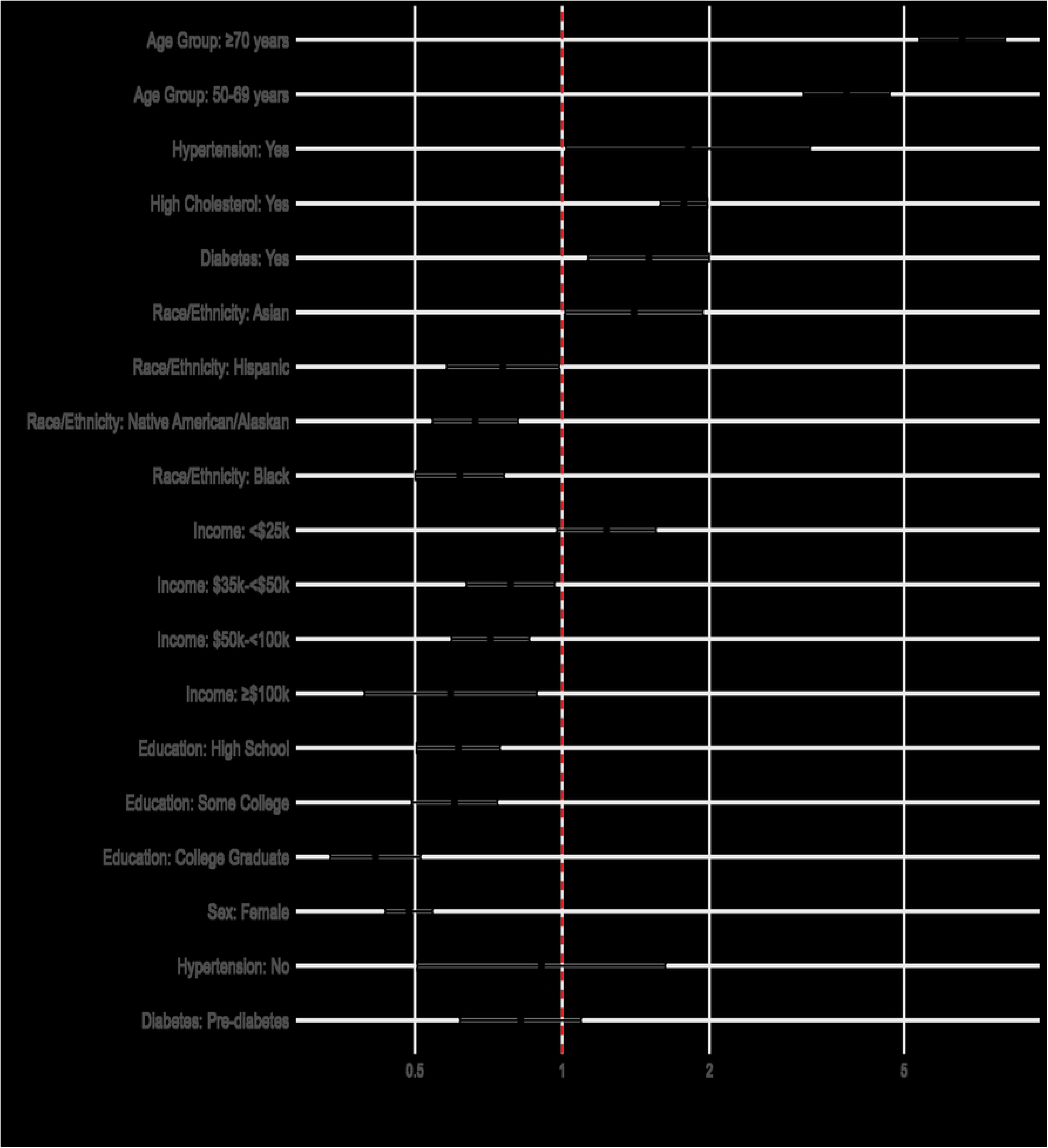
Odds ratios for heart attack risk in the final model. The forest plots display adjusted odds ratios with 95% confidence intervals from the fully adjusted Stage 4 model. The vertical line at 1.0 represents no association (null value). The points to the right (OR>1.0) indicate increased heart attack risk, while the points to the left (OR<1.0) indicate protective associations. The variables are grouped by demographic, socioeconomic, and clinical categories.

**Table 2.** Odds Ratios for Heart Attack Risk in Final Model (Stage 4).

| **Variable** | **Odds Ratio**<br>**(95% CI)** | **P-**<br>**value** |
| --- | --- | --- |
| (Intercept) | 0.03 (0.01-0.06) | <0.001 |
| Sex: Female | 0.49 (0.43-0.54) | <0.001 |
| Age Group: 50-69 years | 3.82 (3.11-4.68) | <0.001 |
| Age Group: $\geq 70$ years | 6.58 (5.37-8.06) | <0.001 |
| Race/Ethnicity: Black | 0.62 (0.50-0.76) | <0.001 |
| Race/Ethnicity: Hispanic | 0.76 (0.58-0.99) | 0.039 |
| Race/Ethnicity: Asian | 1.40 (1.01-1.94) | 0.041 |
| Race/Ethnicity: Native American/Alaskan | 0.66 (0.54-0.81) | <0.001 |
| Income: \$35k-<50k | 0.78 (0.64-0.97) | 0.022 |
| Income: \$50k-<100k | 0.71 (0.59-0.86) | <0.001 |
| Income: <\$25k | 1.23 (0.98-1.56) | 0.08 |
| Income: ≥\$100k | 0.59 (0.39-0.89) | 0.011 |
| Education: High School | 0.61 (0.50-0.75) | <0.001 |
| Education: Some College | 0.60 (0.49-0.74) | <0.001 |
| Education: College Graduate | 0.42 (0.34-0.51) | <0.001 |
| Diabetes: Pre-diabetes | 0.82 (0.62-1.09) | 0.178 |
| Diabetes: Yes | 1.50 (1.13-2.00) | 0.005 |
| Hypertension: No | 0.91 (0.51-1.63) | 0.742 |
| Hypertension: Yes | 1.81 (1.02-3.22) | 0.043 |
| High Cholesterol: Yes | 1.77 (1.59-1.98) | <0.001 |

Note: Analysis based on a random sample of 100,000 observations
from the full dataset.
Note: The analysis is based on the fully adjusted multivariable logistic regression model. The values $>1.0$ indicate increased risk, while the values $<1.0$ indicate protective associations. The reference groups: Male (Sex), 18-49 years (Age), White (Race/Ethnicity), \$25-<35k (Income), Less than High School (Education), No (Diabetes), Borderline (Hypertension), No (High Cholesterol).

The female sex was associated with significantly lower risk (OR=0.49, 95% CI: 0.43- 0.54, p<0.001). Race/ethnicity was also showed significant associations with Black (OR=0.62, 95% CI: 0.50-0.76), Hispanic (OR=0.76, 95% CI: 0.58-0.99), and Native American/Alaskan (OR=0.66, 95% CI: 0.54-0.81). These adults were noted to have a lower risk compared to White adults, after adjusting for other factors. However, Asian adults showed an increased risk (OR=1.40, 95% CI: 1.01-1.94, p=0.041).

The socioeconomic factors in the final model remained consistent, demonstrating significance, even after adjustment for clinical factors. The consistency in these findings demonstrate that higher education was consistently associated with lower risk for heart attack, where college graduates showed the strongest protective factor with an OR=0.42, 95% CI: 0.34- 0.51, and p<0.001. Similarly, this trend continued to follow, where individuals with higher income showed a protective gradient, with income of $100,000 or more were associated with lower risk for a heart attack, with an (OR=0.59, 95% CI: 0.39-0.89, p=0.011).

The clinical factors, such as high cholesterol, showed the strongest association with heart attack risk (OR=1.77, 95% CI: 1.59-1.98, p<0.001), which was then followed by hypertension (OR=1.81, 95% CI: 1.02-3.22, p=0.043) and diabetes (OR=1.50, 95% CI: 1.13-2.00, p=0.005).

However, diabetes was stratified into pre-diabetes, which did not show a significant association with heart attack risk (OR=0.82, 95% CI: 0.62-1.09, p=0.178).

### 3.5 Associations Remaining Significant After Adjustment

The full multivariable adjusted model (Stage 4 model) demonstrated important associations, which retained their statistical significance. There were also other associations that were attenuated and became non-significant. The strongest independent predictor for heart attack remains age, with its’ association maintaining robust significance after adjustment (OR for ≥70 years = 6.58, 95% CI: 5.37-8.06, p<0.001; OR for 50-69 years = 3.82, 95% CI: 3.11-4.68, p<0.001). The clinical risk factors significant independent associations persisted after adjusting for demographic and other comorbidities. The results noted that high cholesterol (OR=1.77, 95% CI: 1.59-1.98, p<0.001), hypertension (OR=1.81, 95% CI: 1.02-3.22, p=0.043), and diabetes (OR=1.50, 95% CI: 1.13-2.00, p=0.005) all remained significant predictors in this model.

However, after adjustment, pre-diabetes became non-significant, suggesting their effects were possibly managed through other variables in the model (OR=0.82, 95% CI: 0.62-1.09, p=0.178).

The demographic factors maintained their protective associations, even after accounting for clinical factors, which include the female sex (OR=0.49, 95% CI: 0.43-0.54, p<0.001) and college education (OR=0.42, 95% CI: 0.34-0.51, p<0.001). Similarly, participants who reported high income kept their protective association (≥$100k: OR=0.59, 95% CI: 0.39-0.89, p=0.011). This analysis demonstrates significant indicators that are independent of other variables, like warning lights on a patient’s health. The variables, age, clinical factors (cholesterol, hypertension, diabetes), and specific demographic characteristics maintained their predictive power even when adjusting for overlapping health influences. This persistence suggests these factors have independent effects on heart attack risk that are not fully explained by other variables in the model.

### 3.6 Model Performance and Validation

#### Staged Model Performance

The staged model performance was important to measure because the model discrimination improved with sequentially adding clinical risk factors (Table 3, Figure 3). The Stage 1 model focused on demographic factors only, which achieved an AUC of 0.763, showing fair discrimination with a 95% CI, 0.763-0.763. Stage 2 added diabetes status to the model, which improved model discrimination as well, achieving an AUC of 0.780, with 95% CI, 0.778- 0.782. The Stage 3 model added hypertension to the model, improving discrimination, achieving an AUC of 0.794, with a 95% CI, 0.793-0.794, showing an increasing trend in the model performance. The last stage, Stage 4, added cholesterol status to the model, showing an AUC of 0.800 with a 95% CI, 0.800-0.800, which demonstrated good model discrimination with a continued trend increase. The Staging approach demonstrates that as these risk factors were added to the model, the model because more accurate. The model had a calibration assessment (Figure 4), which demonstrated how the predicted probabilities and observed risk across all stages aligned excellently, with the Stage 4 model showing a strong calibration (Integrated Calibration Index = 0.009).

**Fig 3.**
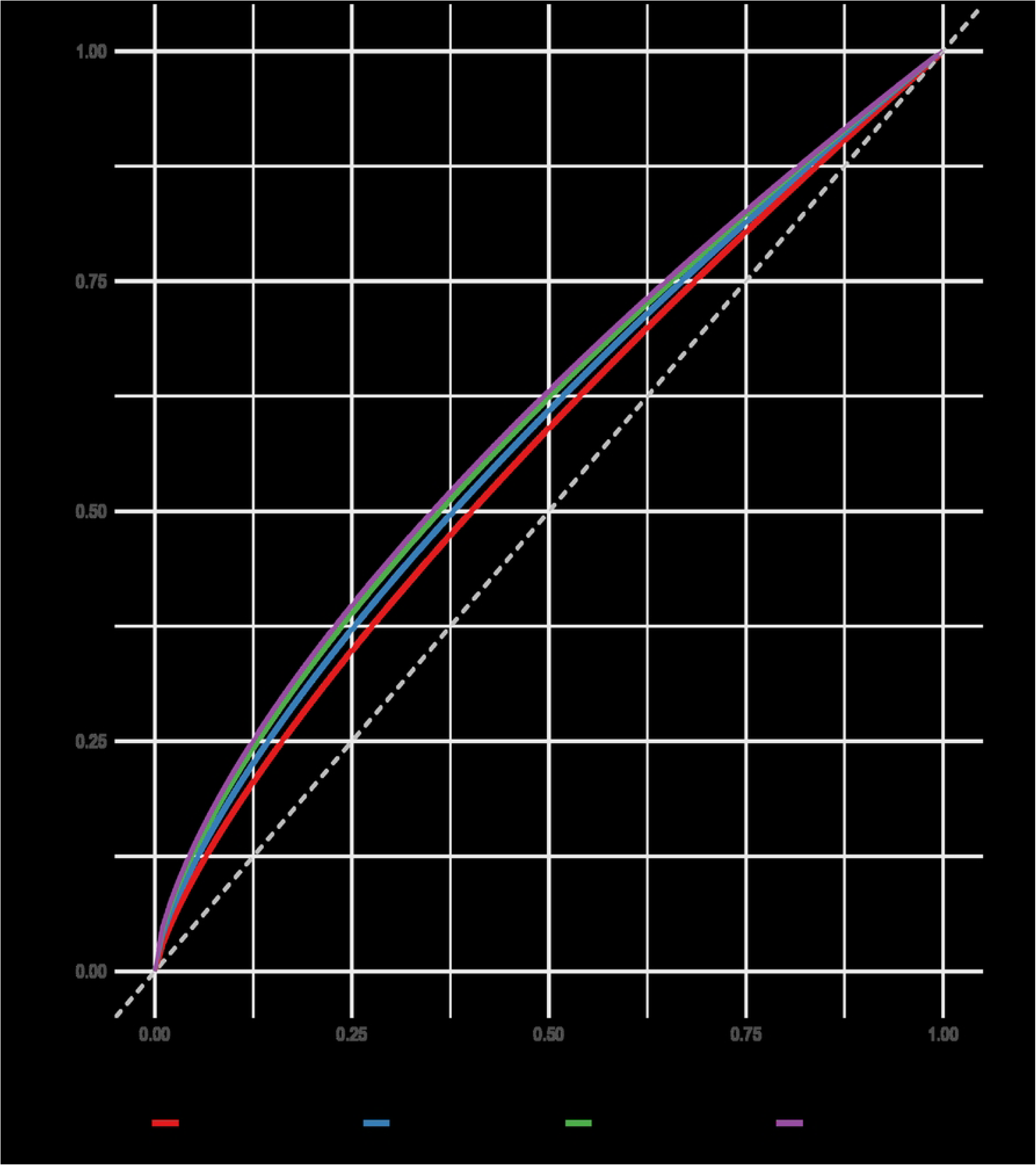
ROC Curves for Sequential Model Stages. The curves display discrimination ability for each modeling stage: Stage 1 (demographics only, AUC=0.763), Stage 2 (adding diabetes, AUC=0.780), Stage 3 (adding hypertension, AUC=0.794), and Stage 4 (adding cholesterol, AUC=0.800). The x-axis represents 1-specificity (false positive rate), and the y-axis represents sensitivity (true positive rate). The diagonal reference line represents chance performance (AUC=0.5).

**Fig 4.**
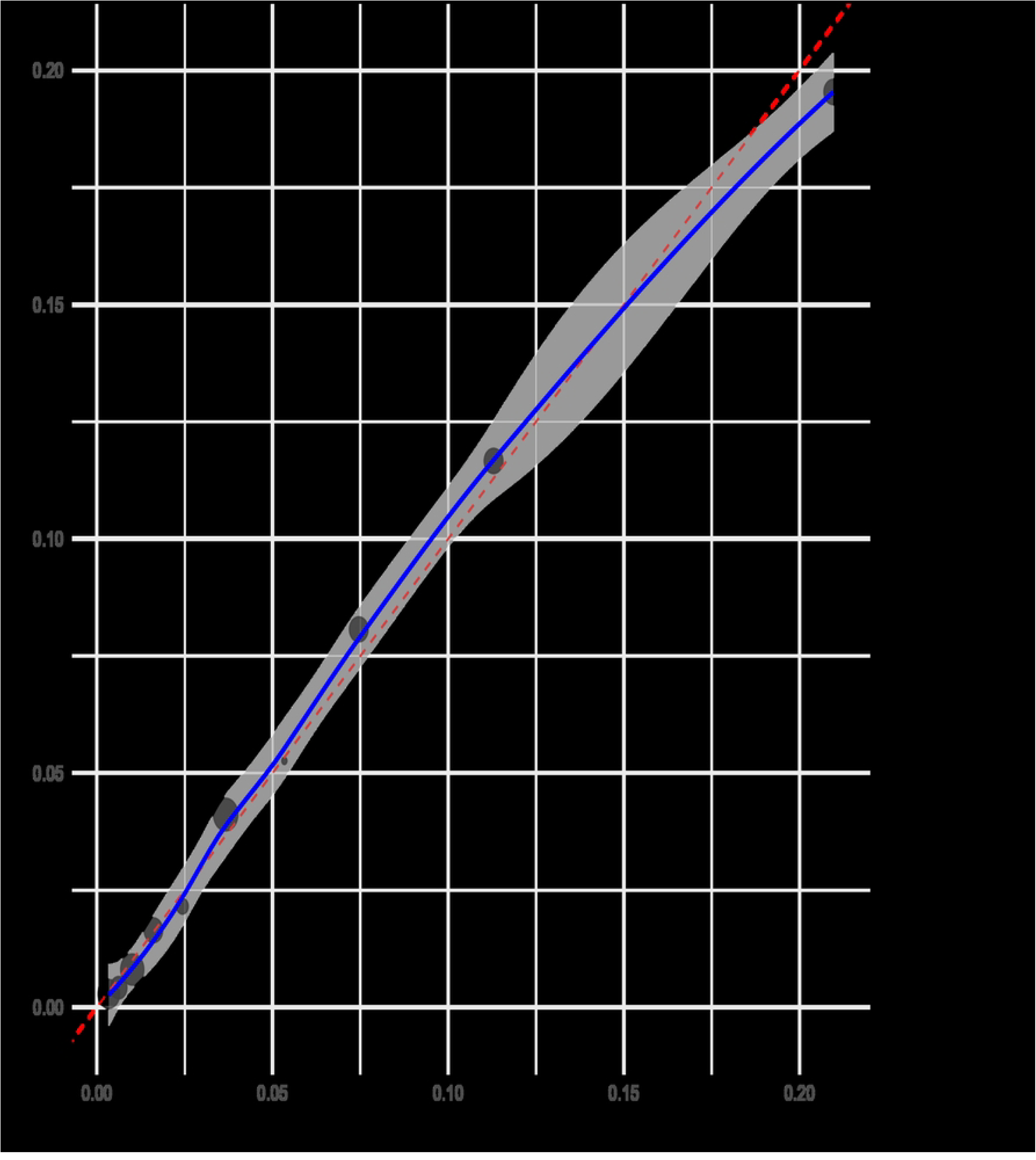
Calibration Plot for Final Model. Calibration plot for the final prediction model. This plot assesses agreement between predicted probabilities (x-axis) and observed outcomes (y-axis). The diagonal line represents perfect calibration. Points show predicted versus observed heart attack rates across deciles of predicted risk, with vertical lines representing 95% confidence intervals. Integrated Calibration Index=0.009 indicates excellent calibration.

**Table 3.** Model Performance Across Progressive Stages.

| Stage | Variables Included | AUC-ROC (95% CI) | Integrated Calibration Index (ICI) |
| --- | --- | --- | --- |
| 1 | Demographics | 0.763 (0.763-0.763) | 0.013 |
| 2 | + Diabetes | 0.780 (0.778-0.782) | 0.011 |
| 3 | + Hypertension | 0.794 (0.793-0.794) | 0.010 |
| 4 | + Cholesterol | 0.800 (0.800-0.800) | 0.009 |

Note: AUC-ROC = Area Under the Receiver Operating Characteristic Curve; ICI = Integrated Calibration Index. Stage 1 includes demographic factors only; Stage 2 adds diabetes; Stage 3 adds hypertension; Stage 4 adds cholesterol status.

#### Sensitivity Analyses

The final model performance was consistent across sex groups, with similar discrimination for males (AUC=0.794) and females (AUC=0.787). The age-stratification analysis showed moderate discrimination within each age group, with participants between the ages of 18-49 years (AUC=0.767), 50-69 years (AUC=0.751), and ≥70 years (AUC=0.683). The variable importance analysis (Figure 5) identified age as the most critical predictor for heart attack risk, with its removal, leading to the largest decrease in AUC (-0.058), followed by cholesterol status (-0.013), sex (-0.009), and then diabetes status (-0.007). The two variables, education and income, contributed modestly to model discrimination (AUC decrease of -0.006 and -0.004, respectively).

**Fig 5.**
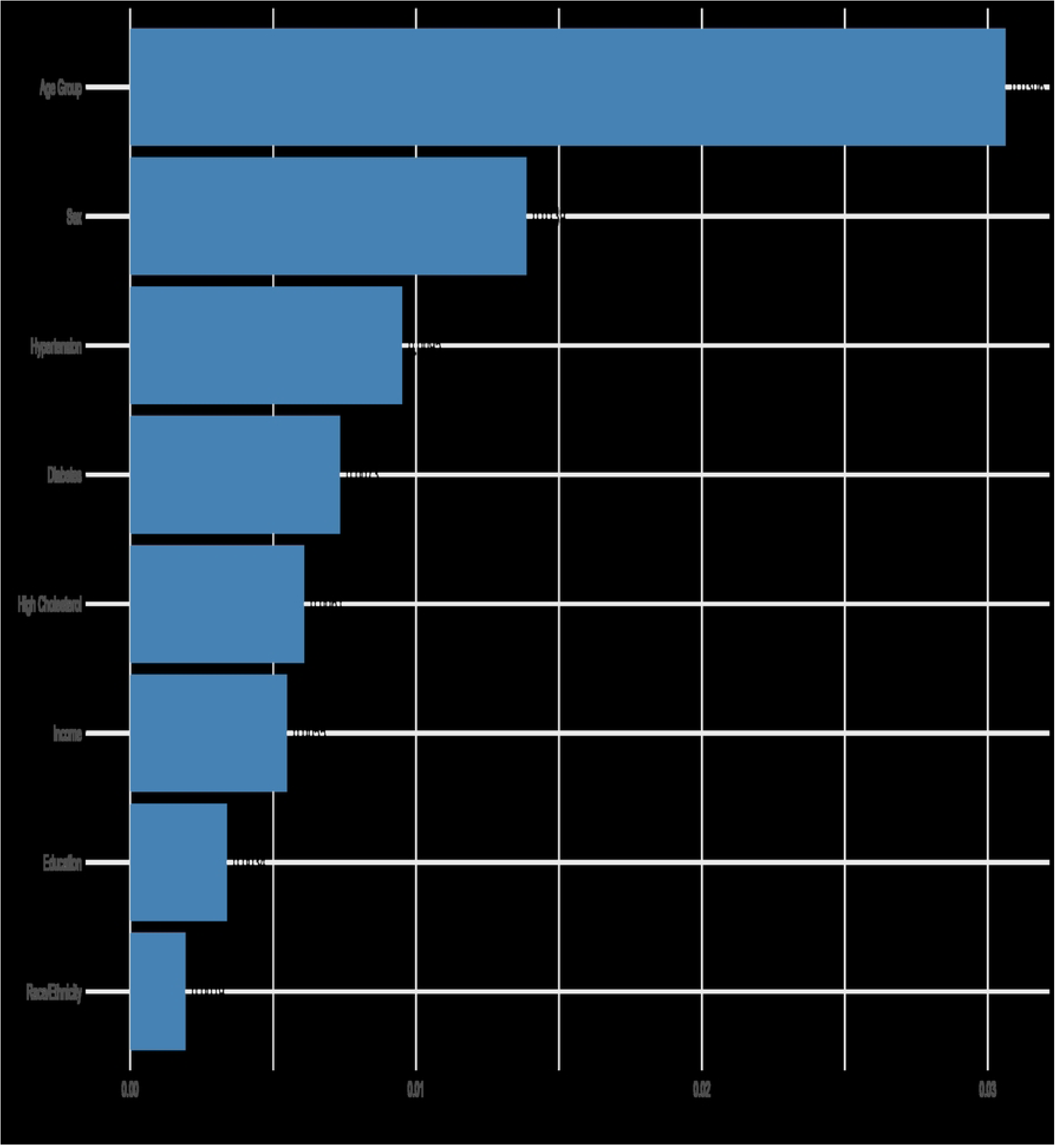
Variable Importance Analysis. Variable importance in heart attack risk prediction. This plot shows the reduction in Area Under the Curve (AUC) when each predictor variable is removed from the final model, quantifying each variable’s contribution to model discrimination. Larger values indicate greater importance to predictive performance. Age Group demonstrates the strongest contribution (AUC reduction of 0.0306), followed by Sex (0.0139), Hypertension (0.0095), and Diabetes (0.0073).

## Discussion

### 4.1 Principal Findings

As hypothesized, demographic factors such as age, sex, socioeconomic status, and education established a foundation discrimination for heart attack risk. The staged approach of clinical factors, such as diabetes, hypertension, and high cholesterol, yielded incremental improvements in the predictive performance, confirming this study’s hypothesis. The principal findings of this research were to develop and validate a heart attack prediction model using nationally representative survey data (BRFSS) by deploying a staged approach that integrates demographic and clinical risk factors. The findings demonstrated noteworthy discrimination (AUC=0.800), and calibration can be achieved with a parsimonious model, which included age, sex, race/ethnicity, socioeconomic indicator, and three key clinical risk factors (diabetes, hypertension, and high cholesterol).

The staged approach established strong predictive value from demographic factors (AUC=0.763), which improved when adding clinical factors incrementally. The findings suggested that demographics can provide meaningful insights on heart attack risk, especially when clinical data is absent. The additional self-reported clinical factors yielded a more comprehensive and discriminating model that can be seen as the stages increased from baseline to Stage 4 of the model. Our findings directly addressed the four research questions posed at the outset. Regarding which populations have the highest heart attack burden, we identified older adults (≥70 years: 10.9%), men (6.7%), those with less education (less than high school: 10.2%), lower income (<$25k: 8.8%), and those with diabetes (12.9%), hypertension (9.6%), or high cholesterol (8.8%) as bearing the highest burden.

The analysis revealed significant health disparities across all demographic categories studied (p<0.001), with the most striking differences emerging across age, income brackets, and education levels. When examining which factors posed the greatest risk for heart attacks, this study found that individuals aged 70 years and older faced the highest risk compared to young adults (OR=6.58). This was followed by other major risk factors, such as hypertension (OR=1.81), high cholesterol (OR=1.77), and diabetes (OR=1.50). Even after statistical adjustments, with all the covariates, these clinical factors remained significant. Similarly, there were key demographic factors including age, sex, and education, that maintained their predictive value in the final model. However, some factors like pre-diabetes, lost their statistical significance once other variables were controlled.

### 4.2 Comparison with Prior Literature

The findings in this research align with previous cardiovascular risk prediction models while offering several unique contributions, which can be important when considering heart attack risk. The observed associations between traditional risk factors and heart attack risk factors are consistent and have been well established with cardiovascular predictive tools such as the Framingham Risk Score[14][15] and the Pooled Cohort Equations[16][17]. However, these models typically rely on laboratory values and clinical measurements, while our model achieves comparable discrimination using only self-reported information by the participants. The strong protective association of higher education (OR=0.42) for college graduates was particularly noteworthy and consistent with the continuously growing evidence regarding socioeconomic factors as independent cardiovascular risk factors. These findings align with the findings from the REGARDS study[18][19] and highlight the importance of considering social determinants of health when actively assessing participants for heart attack risk.

The model presented in this research demonstrates strong discrimination with an AUC of 0.800, which compares favorably with existing predictive tools, showing strength in these results. The 2008 study that utilized the Framingham Risk Score reported AUCs of 0.763 in men and 0.793 in women[20], while the 2013 ACC/AHA Pooled Cohort Equation achieved AUCs between 0.71 and 0.82 across various populations[21]. These, align with the results of this research, show not only comparable discrimination, but also the importance of evaluating social determinants of health in clinical presentation. Notably, the model in this research achieves performance without requiring laboratory measurements or detailed clinical assessments, which is important considering its utility for population-level screening.

### 4.3 Clinical and Public Health Implications

The staged modeling approach demonstrates that demographic information can provide substantial risk stratification and aid in assessing heart attack risk assessment (AUC=0.763). This is an important implication for providers in resource-limited settings, where clinical data may not be readily or quickly available or accessible. The model could serve as a simple screening tool to identify individuals for more intensive assessments or targeted interventions, such as moving forward with laboratory measurements or detailed clinical assessments.

The incremental value of each clinical factor in the staging model (Stage 1-4) suggests potential prioritization for risk assessments, showing that cholesterol status provided the largest improvement in discrimination among other clinical factors. This is supported with current guidelines, emphasizing that cholesterol management for primary prevention is important[22].

Diabetes and hypertension are also significant independent contributors that underscore the importance of comprehensive risk factor management.

The persistent protective effect of higher education after adjusting for clinical factors is essential, as this highlights the need for providers to consider social determinants of health in cardiovascular risk assessments and intervention planning. The findings of this research suggest that population-level approaches, such as addressing educational disparities, could complement the individual clinical interventions, which can aid in the reduction of cardiovascular burden.

The fully adjusted model demonstrates some disparities that are important to discuss, as Black, Hispanic, and Native American/Alaskan participants had significantly lower odds ratios of self-reported heart attack compared to White participants. These findings are counter to national mortality statistics and may reflect underdiagnosis, underreporting, or survivor bias in self-reported survey data, as well as potential residual confounding. These results highlight the need for cautious interpretation of self-reported outcomes in population surveys and underscore the importance of improving healthcare access and diagnostic equity, thus circling back to the importance of social determinants of health.

#### 4.3.1 Racial Disparities and Mortality Paradox

Our model showed lower odds of self-reported heart attacks among Black (OR=0.62), Hispanic (OR=0.76), and Native American/Alaskan (OR=0.66) participants compared to White individuals. This contrasts sharply with a circulation study on cardiovascular mortality in 2019 was higher for Black women (rate ratio, 1.32 [95% CI, 1.30–1.33]) as well as for Black men (rate ratio, 1.33 [95% CI, 1.32–1.34]), compared with their respective White participants^23^. These disparities likely reflect systemic underdiagnosis in self-reported surveys rather than biological protection. Underserved populations are no stranger to facing barriers to cardiovascular care.

These barriers typically consist of delayed diagnoses, limited access to confirmatory testing, and higher competing mortality risks from untreated comorbidities^11, 24^. These structural inequities may be the result of lower detection rates of heart attacks in populations that reported their findings to BRFSS.

### 4.4 Strengths and Limitations

**Strengths:** This study has several notable strengths that help demonstrate the utility of this research. The first strength is the nationally representative dataset, which promotes generalizability of these findings. The large sample from BRFSS allows this research to randomly sample 100,000 participants out of 430,755, providing a robust outcome. Secondly, the staged approach demonstrates incremental value of each risk factor group, which demonstrates the effects of risk factors as they continue to grow. Thirdly, this research used rigorous methodologies for managing missing data by using multiple imputations for more precise results. Fourthly, the research used comprehensive validation methods, including model discrimination, calibration, and a sensitivity analysis to demonstrate model robustness. Lastly, the parsimonious nature of this model enhances its practice utility for population-level screening.

**Limitations:** There were several limitations that warrant consideration for this model as well. The first limitation is cross-sectional research design, which is the nature of BRFSS data, and this precludes assessment of incident heart attacks, limiting the model to prevalent cases. The cross-sectional by design prevents causal inference, therefore, the strong age gradient (OR=6.58 for ≥70y vs 18-49y) aligns with other longitudinal studies evaluating CVD risk[25][26]. These findings suggest our model captures fundamental aging-related risk pathways. However, survivor bias may diminish observed associations, particularly in older adults who survived prior CVD events. The second limitation in this study is the reliance on self-reported data, which may introduce recall bias into the study, even though previous validation studies have shown reasonable accuracy for major chronic conditions in BRFSS[27][28].

There were other limitations noted as well, such as reporting bias may differ across different demographic groups. The limitation with this bias is the possibility that it may lead to potentially underreporting among racial/ethnic minorities and those who come from a lower socioeconomic status. This is likely associated with disparities in healthcare access, diagnosis rates, and health literacy. This limitation and differential reporting could partially explain the unexpected protective associations observed for certain racial/ethnic groups after adjustment. The third limitation focused on clinical measurements and biomarkers, which prevent this research from capturing subclinical disease and more nuanced risk profiles. Fourthly, this model demonstrates strong discrimination and calibration in the development dataset, and additionally, external validation in independent cohorts would strengthen the confidence of these findings and its generalizability. Lastly, this research does not account for medication use or treatment effects, which may influence associations between risk factors and outcomes.

### 4.5 Future Research

There are several directions for future research to emerge from these findings, all which can improve the outcome of heart attack risk assessments. The first suggestion is external validation, which can be used in prospective cohorts, as this would strengthen evidence for the model’s predictive validity for incident events. Second, this can be used to compare with other models that incorporate laboratory values, which could help quantify the potential trade-offs between a simplified approach and predictive power. The third suggestion is integrating additional risk factors, which include physical activity, dietary habits, and psychosocial factors. These additional factors may improve model discrimination through similar staging approaches. Fourth, this research sets a future for the development of risk calculators or simplified scoring systems that can be used in clinical settings with limited resources. Lastly, future researchers can evaluate model performance across diverse populations, particularly populations with underrepresented minorities, which can ensure a more equitable heart attack risk assessment.

## Conclusion

This study developed and validated a heart attack prediction model using a staged approach (Stages 1-4), which integrated demographic and clinical risk factors from nationally representative survey data (BRFSS). The final model achieved noteworthy discrimination (AUC=0.800) and calibration, which demonstrated that an accurate cardiovascular risk assessment can be accomplished using self-reported information, without the use of laboratory values or detailed clinical measurements and data. The staged approach in this research demonstrated substantial predictive value from just demographic factors, and then incrementally improved as clinical factors were added to the assessment, such as diabetes, hypertension, and cholesterol status. It is worth noting that age emerged as the strongest predictor in this model, followed by clinical factors and sex, However, socioeconomic indicators maintained independent protective associations even after adjusting for traditional risk factors. The model performed consistently across sex groups, with comparable discrimination for both male and female participants.

These findings have important implications for population-level health and clinical practice. The model could serve as an efficient screening tool to identify high-risk individuals in settings where comprehensive clinical assessments are impractical, while the staged approach informs efficient risk stratification strategies for providers. It is important to also note that the persistent protective factor of higher education highlights the importance of providers addressing social determinants of health alongside clinical risk factors when considering a comprehensive cardiovascular disease prevention plan. Future research should focus on external validation in prospective cohort studies, as well as compare this with laboratory-based models, and potentially lead to developing simplified risk calculators for clinical implementation. Accessible and accurate risk prediction tools are essential in assessing heart attack risk and they must be effective at prevention and control strategies because cardiovascular disease remains a leading cause of morbidity and mortality worldwide. The parsimonious model used in this research balances predictive performance with practical utility, offering a valuable tool for population-level cardiovascular risk assessments, particularly heart attack risk.

## Data Availability

All data underlying the findings described in this manuscript are fully available without restriction. The BRFSS 2023 data are publicly available from the Centers for Disease Control and Prevention (CDC) at: https://www.cdc.gov/brfss/annual_data/annual_2023.html. The analytical code used in this study is available on GitHub at: https://github.com/adamschomer/HeartAttackRisk

https://www.cdc.gov/brfss/annual_data/annual_2023.html

## References

1. Lewis C. Heart disease remains leading cause of death as key health risk factors continue to rise [Internet]. American Heart Association. 2025 [cited 2025 May 31]. Available from: https://newsroom.heart.org/news/heart-disease-remains-leading-cause-of-death-as-key-health-risk-factors-continue-to-rise

2. Jain V, Al Rifai M, Khan SU, Kalra A, Rodriguez F, Samad Z, et al. Association Between Social Vulnerability Index and Cardiovascular Disease: A Behavioral Risk Factor Surveillance System Study. J Am Heart Assoc. 2022;11(15):e024414. doi:10.1161/JAHA.121.024414

3. Centers for Disease Control and Prevention. Heart disease facts [Internet]. Atlanta: CDC; 2024 [cited 2025 Jun 1]. Available from: https://www.cdc.gov/heart-disease/data-research/facts-stats/index.html

4. Singh R, Javed Z, Yahya T, Valero-Elizondo J, Acquah I, Hyder AA, et al. Community and Social Context: An Important Social Determinant of Cardiovascular Disease. Methodist DeBakey Cardiovasc J. 2021;17(4):15–27. doi:10.14797/mdcvj.846

5. Powell-Wiley TM, Baumer Y, Baah FO, Baez AS, Farmer N, Mahlobo CT, et al. Social Determinants of Cardiovascular Disease. Circ Res. 2022;130(5):782–99. doi:10.1161/CIRCRESAHA.121.319811

6. Schultz WM, Kelli HM, Lisko JC, Varghese T, Shen J, Sandesara P, et al. Socioeconomic Status and Cardiovascular Outcomes: Challenges and Interventions. Circulation. 2018;137(20):2166-78. doi:10.1161/CIRCULATIONAHA.117.029652

7. Abdalla SM, Rosenberg SB, Maani N, Contreras CM, Yu S, Galea S. Income, education, and the clustering of risk in cardiovascular disease in the US, 1999–2018: An observational study. Lancet Reg Health Am. 2018;0(0):101039. doi:10.1016/j.lana.2025.101039

8. Rodgers JL, Jones J, Bolleddu SI, Vanthenapalli S, Rodgers LE, Shah K, et al. Cardiovascular Risks Associated with Gender and Aging. J Cardiovasc Dev Dis. 2019;6(2):19. doi:10.3390/jcdd6020019

9. McClellan M, Brown N, Califf RM, Warner JJ. Call to Action: Urgent Challenges in Cardiovascular Disease: A Presidential Advisory From the American Heart Association. Circulation. 2019;139(9):e0652. doi:10.1161/CIR.0000000000000652

10. Mazimba S, Peterson PN. JAHA Spotlight on Racial and Ethnic Disparities in Cardiovascular Disease. J Am Heart Assoc. 2021;10(17):e023650. doi:10.1161/JAHA.121.023650

11. Lindley KJ, Aggarwal NR, Briller JE, Davis MB, Douglass P, Epps KC, et al. Socioeconomic Determinants of Health and Cardiovascular Outcomes in Women: JACC Review Topic of the Week. J Am Coll Cardiol. 2021;78(19):1919–29. doi:10.1016/j.jacc.2021.09.011

12. Spruill TM, Butler MJ, Thomas SJ, Tajeu GS, Kalinowski J, Castañeda SF, et al. Association Between High Perceived Stress Over Time and Incident Hypertension in Black Adults: Findings From the Jackson Heart Study. J Am Heart Assoc. 2019;8(21):e012139. doi:10.1161/JAHA.119.012139

13. Centers for Disease Control and Prevention. Chronic Kidney Disease, Diabetes, and Heart Disease [Internet]. Atlanta: CDC; 2024 [cited 2025 Jun 5]. Available from: https://www.cdc.gov/kidney-disease/risk-factors/link-between-diabetes-and-heart-disease.html

14. Luiz AA de MJ, Silva S, Cardoso C, Nascimento S, Aloisio F, Luiz G, et al. Framingham score adapted: A valid alternative for estimating cardiovascular risk in epidemiological studies. BMC Cardiovasc Disord. 2025;25(1):e12345. doi:10.1186/s12872-025-04579-x

15. Zeynalova S, Bucksch K, Scholz M, Yahiaoui-Doktor M, Gross M, Löffler M, et al. Monocyte subtype counts are associated with 10-year cardiovascular disease risk as determined by the Framingham Risk Score among subjects of the LIFE-Adult study. PLoS One. 2021;16(3):e0247480. doi:10.1371/journal.pone.0247480

16. Shetty NS, Mokshad G, Patel N, Nehal V, Li P, Arora G, et al. PREVENT and Pooled Cohort Equations in Mortality Risk Prediction. JACC Adv. 2024;3(12):101372. doi:10.1016/j.jacadv.2024.101372

17. Zhou H, Zhang Y, Zhou MM, Choi SK, Reynolds K, Harrison TN, et al. Evaluation and Comparison of the PREVENT and Pooled Cohort Equations for 10-Year Atherosclerotic Cardiovascular Risk Prediction. J Am Heart Assoc. 2025;14(4):e039454. doi:10.1161/JAHA.124.039454

18. Kaplan RM, Howard VJ, Safford MM, Howard G. Educational attainment and longevity: results from the REGARDS U.S. national cohort study of blacks and whites. Ann Epidemiol. 2015;25(5):323–8. doi:10.1016/j.annepidem.2015.01.017

19. Lewis MW, Khodneva Y, Redmond N, Durant RW, Judd SE, Wilkinson LL, et al. The impact of the combination of income and education on the incidence of coronary heart disease in the prospective Reasons for Geographic and Racial Differences in Stroke (REGARDS) cohort study. BMC Public Health. 2015;15(1):e12345. doi:10.1186/s12889-015-2630-4

20. D’Agostino RB, Vasan RS, Pencina MJ, Wolf PA, Cobain M, Massaro JM, et al. General Cardiovascular Risk Profile for Use in Primary Care. Circulation. 2008;117(6):743–53. doi:10.1161/CIRCULATIONAHA.107.699579

21. Goff DC, Lloyd-Jones DM, Bennett G, Coady S, D’Agostino RB, Gibbons R, et al. 2013 ACC/AHA Guideline on the Assessment of Cardiovascular Risk. Circulation. 2013;129(25 suppl 2):S49-73. doi:10.1161/01.cir.0000437741.48606.9

22. Grundy SM. 2018 AHA/ACC/AACVPR/AAPA/ABC/ACPM/ADA/AGS/APhA/ASPC/NLA/PCNA Guideline on the Management of Blood Cholesterol. Circulation. 2019;139(25). doi:10.1161/CIR.0000000000000625

22. Kyalwazi AN, Loccoh EC, Brewer LC, Ofili EO, Xu J, Song Y, et al. Disparities in Cardiovascular Mortality Between Black and White Adults in the United States, 1999 to 2019. Circulation. 2022;146(3):211-28. doi:10.1161/CIRCULATIONAHA.122.060199

23. Faugno E, Galbraith AA, Walsh K, Maglione PJ, Farmer JR, Ong MS. Experiences with diagnostic delay among underserved racial and ethnic patients: A systematic review of the qualitative literature. BMJ Qual Saf. 2024;34(3):bmjqs-2024-017506. doi:10.1136/bmjqs-2024-017506

24. Kaneko H, Yano Y, Okada A, Itoh H, Suzuki Y, Yokota I, et al. Age-Dependent Association Between Modifiable Risk Factors and Incident Cardiovascular Disease. J Am Heart Assoc. 2023;12(2):e027684. doi:10.1161/JAHA.122.027684

25. Lind L, Sundström J, Ärnlöv J, Ingelsson M, Henry A, Lumbers RT, et al. Life-Time Covariation of Major Cardiovascular Diseases. Circ Genom Precis Med. 2021;14(2):e002963. doi:10.1161/CIRCGEN.120.002963

26. Watson KB, Wiltz JL, Nhim K, Kaufmann RB, Thomas CW, Greenlund KJ. Trends in Multiple Chronic Conditions Among US Adults, By Life Stage, Behavioral Risk Factor Surveillance System, 2013–2023. Prev Chronic Dis. 2025;22:240539. doi:10.5888/pcd22.240539

27. Leal R, Spencer-Hwang R, Beeson L, Paalani M, Dos Santos H. Lifestyle Factors and Heart Health: Exploring Effect Modification Using Behavioral Risk Factor Surveillance System Survey Data. Am J Lifestyle Med. 2024 Jan 19. doi:10.1177/15598276241226930

